# Shared Genetics of Hypertension and Preeclampsia Converges on Immune Regulation

**DOI:** 10.64898/2026.05.05.26352450

**Authors:** Mohammed A. Farahat, Malak Abbas, Mileati Melese, Amadou Gaye¹

## Abstract

**Background:** Hypertension and preeclampsia are clinically distinct, yet biologically related conditions characterized by vascular dysfunction and elevated cardiovascular risk. Although genome-wide association studies (GWAS) have identified loci associated with blood pressure traits and preeclampsia, the functional mechanisms linking shared variants to gene regulation and clinical phenotypes remain unclear.

**Methods:** We integrated GWAS summary statistics for hypertension, systolic blood pressure (SBP), diastolic blood pressure (DBP), and preeclampsia to identify shared variants (p ≤ 1×10⁻⁴). Cis-expression quantitative trait loci (eQTL) analyses were performed in whole blood using RNA-seq data from 180 African American women. Significant associations (FDR ≤ 0.05) were evaluated for replication across vascular, metabolic, and endocrine tissues in the Genotype-Tissue Expression (GTEx) project. Associations between gene expression and blood pressure traits were also assessed.

**Results:** We identified 4,792 shared GWAS variants, of which 4,663 were tested in eQTL analyses, yielding 1,837 significant variant-gene associations across 78 genes. Replication in GTEx confirmed 645 associations involving 24 genes, many showing cross-tissue regulatory effects. Three genes (C4B, HLA-C, and HLA-DQB1) demonstrated convergent evidence across GWAS, gene regulation, and expression-trait analyses. C4B expression was positively associated with hypertension and SBP, while HLA-C showed consistent negative associations with hypertension, SBP, and DBP. HLA-DQB1 expression was specifically associated with DBP, suggesting trait-specific effects.

**Conclusions:** These findings highlight immune-related pathways as key mediators linking hypertension and preeclampsia. Integrating genetic, transcriptomic, and phenotypic data provides a framework for identifying functionally relevant loci and advancing mechanistic insights into cardiometabolic and pregnancy-related disorders.

**GRAPHICAL ABSTRACT:** 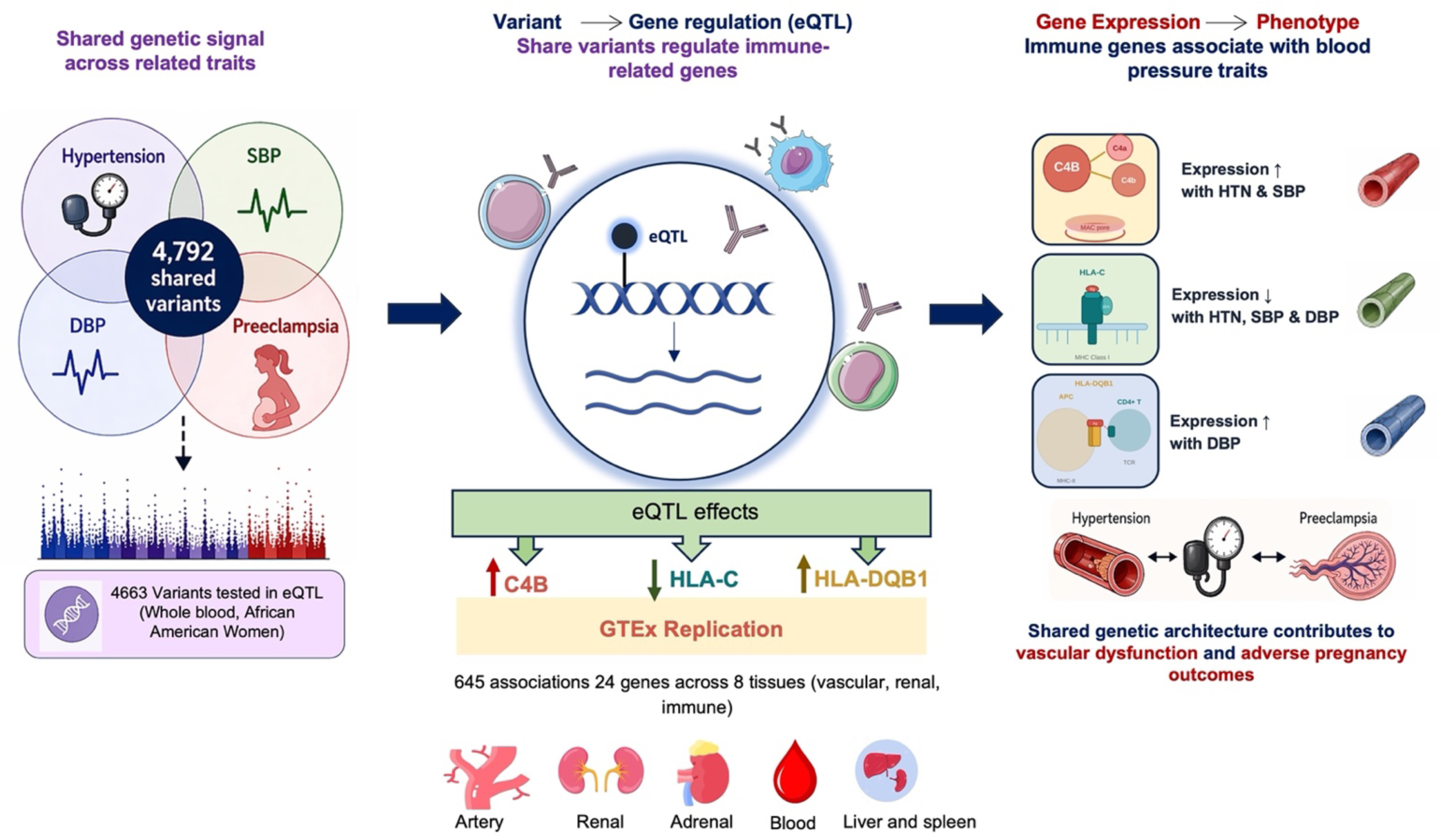

**Shared genetic variants across hypertension, blood pressure traits, and preeclampsia converge on immune regulatory genes linking gene regulation to clinical phenotypes.**

GWAS summary statistics for hypertension, SBP, DBP, and preeclampsia were intersected to identify 4,792 shared variants, of which 4,663 were tested in cis-eQTL analyses in whole blood from 180 African American women (*left*). Shared variants regulate immune-related genes through cis-eQTL effects, yielding 1,837 associations involving 78 genes (FDR ≤ 0.05). Three convergent genes emerged: C4B (upregulated), HLA-C (downregulated), and HLA-DQB1 (upregulated), with 645 associations involving 24 genes replicated across eight tissues in GTEx (*center*). Expression-trait analyses confirmed that C4B expression was positively associated with hypertension and SBP, HLA-C expression was negatively associated with hypertension, SBP, and DBP, and HLA-DQB1 expression was specifically associated with DBP. These genes implicate complement activation, antigen presentation, and adaptive immunity as shared mechanisms contributing to vascular dysfunction in both hypertension and preeclampsia.

eQTL indicates expression quantitative trait locus; FDR, false discovery rate; GTEx, Genotype-Tissue Expression project; SBP, systolic blood pressure; DBP, diastolic blood pressure; APC, antigen-presenting cell; TCR, T-cell receptor; MHC, major histocompatibility complex.

## INTRODUCTION

Hypertension is a leading contributor to global morbidity and mortality, affecting approximately 1.4 billion of the adult population worldwide and representing a major risk factor for cardiovascular and renal disease^1, 2^. Preeclampsia, a pregnancy-specific hypertensive disorder characterized by new-onset hypertension after 20 weeks of gestation, complicates approximately 3-5% of pregnancies and remains a major cause of maternal and fetal morbidity and mortality globally^3^. Although traditionally considered distinct clinical entities, accumulating evidence suggests that hypertension and preeclampsia are closely related conditions along a shared pathophysiological spectrum. Women who develop preeclampsia have an increased long-term risk of chronic hypertension and cardiovascular disease later in life, supporting the notion of shared underlying mechanisms^4^.

Both hypertension and preeclampsia are complex, multifactorial conditions influenced by genetic, environmental, and behavioral factors. Genome-wide association studies (GWAS) have identified numerous loci associated with blood pressure traits, including systolic blood pressure (SBP), diastolic blood pressure (DBP), and hypertension, as well as a growing number of loci linked to preeclampsia^5–8^. These studies have provided important insights into disease susceptibility and have highlighted biological pathways related to vascular function, renal regulation, immune activation, and metabolic processes^8^. However, most genetic studies have investigated these traits independently, limiting our understanding of the extent to which they share common genetic determinants.

The concept of shared genetic architecture across related traits offers a powerful framework to uncover common biological mechanisms. Given the overlapping clinical features and epidemiological associations between hypertension and preeclampsia, it is plausible that genetic variants contributing to blood pressure regulation may also influence susceptibility to preeclampsia. Yet, systematic efforts to identify and functionally characterize genetic variants shared across these traits remain limited. In particular, there is an important gap in translating cross-trait genetic signals into mechanistic insights, as the functional consequences of many GWAS-identified variants remain poorly understood^8, 9^.

A major challenge in the interpretation of GWAS findings is that most associated variants reside in non-coding regions of the genome, where they are thought to exert regulatory effects on gene expression rather than directly altering protein sequence^10^. Expression quantitative trait loci (eQTL) analyses provide a framework to bridge this gap by linking genetic variation to gene expression levels. By integrating GWAS findings with transcriptomic data, eQTL analyses enable the identification of regulatory variants and their target genes, offering insights into the molecular mechanisms through which genetic risk contributes to disease^11^. Importantly, the effects of regulatory variants are often tissue-specific, underscoring the need to consider biologically relevant tissues when interpreting genetic associations^12^.

In the context of hypertension and preeclampsia, several tissue systems are likely to play key roles, including vascular tissues involved in endothelial function and vascular tone, renal tissues responsible for fluid and electrolyte balance, immune-related compartments reflecting systemic inflammation, and metabolic and endocrine tissues that influence cardiometabolic homeostasis. Understanding how shared genetic variants act across these tissues is essential for revealing the biological pathways linking chronic hypertension and pregnancy-related hypertensive disorders.

In this study, we sought to identify and functionally characterize genetic variants shared across hypertension-related traits, including SBP, DBP, and preeclampsia, by integrating GWAS and transcriptomic data. We first identified trait-specific variants and defined a set of variants shared across all four phenotypes. We then evaluated the regulatory effects of these variants on gene expression using whole-blood transcriptomic data from the Genomics, Environmental Factors, and the Social Determinants of Cardiovascular Disease in Africans Americans Study GENE-FORECAST cohort. Next, we assessed the replication and tissue specificity of the identified eQTL-gene associations across multiple biologically relevant tissues using data from the Genotype-Tissue Expression (GTEx) project^13^. Finally, we examined whether the expression levels of eQTL-target genes are associated with blood pressure traits in the same sample set, providing an additional layer of functional evidence linking genetic regulation to clinical phenotypes.

By focusing on shared genetic architecture and integrating genomic and transcriptomic data, this study aims to move beyond association signals toward a mechanistic understanding of how common genetic variation influences both blood pressure regulation and preeclampsia. These insights may help clarify the biological links between chronic and pregnancy-related hypertension and inform future strategies for risk prediction and therapeutic targeting.

## MATERIALS AND METHODS

### Study Cohort

The GENE-FORECAST study is a population-based research platform designed to generate deep phenotyping data in African American adults through an integrated multi-omics framework. The cohort comprises U.S.-born African American adults aged 21-65 years, recruited primarily from communities in the greater Washington, DC, area using a community-engaged enrollment strategy, as detailed previously^12^. The study protocol was approved by the Institutional Review Boards of the National Institutes of Health and Meharry Medical College. Written informed consent was obtained from all participants prior to enrollment, and all study procedures adhered to institutional and regulatory requirements.

### Genotype Data

Whole-genome sequencing was performed on DNA isolated from EDTA-anticoagulated peripheral blood. DNA concentrations were quantified using the PicoGreen assay. Sequencing libraries were prepared using a PCR-free, small-insert whole-genome workflow, and samples were sequenced as 151-bp paired-end reads on the Illumina NovaSeq 6000 platform, achieving an average depth of 30X. Raw reads were preprocessed using the Broad Institute’s Whole Genome Germline Variant Discovery pipeline. Alignment to the human reference genome (hg38) was performed with the Burrows-Wheeler Aligner (BWA), and variant calling, gVCF aggregation, joint genotyping, and subsequent quality control followed GATK4 best-practice recommendations. Of the participants with WGS data, 180 women also had matched whole-blood transcriptomic profiles generated by mRNA sequencing.

### Transcriptomics Data

Whole-blood transcriptomic profiles were generated using messenger RNA sequencing (mRNA-seq). Total RNA was isolated from stabilized blood tubes with the MagMAX™ RNA extraction kit (Life Technologies, Carlsbad, CA). Sequencing libraries were prepared from total RNA using Illumina TruSeq chemistry to generate indexed cDNA libraries after depletion of ribosomal RNA.

Paired-end sequencing was performed on Illumina HiSeq 2500 and HiSeq 4000 instruments, with each sample sequenced to a minimum depth ≥ 50 million reads. Gene expression quantification followed the workflow implemented by the Broad Institute for GTEx, as described in their publicly available pipeline. Transcripts with low abundance, defined as fewer than 2 counts per million (CPM) in at least three samples, were removed prior to normalization. Expression values were then normalized using the Trimmed Mean of M-values approach, which is well-suited for count-based RNA-seq data³. Principal component analysis was used to identify expression outliers, leading to the exclusion of four transcripts.

After quality-control filtering, 17,947 protein-coding genes were retained for downstream analyses. The eQTL mapping was carried out in the subset of 180 women for whom both WGS and mRNA-seq data were available.

### eQTL Analysis

The analytical workflow comprised three major components (**Figure 1**). We first retrieved summary statistics from large-scale GWAS of hypertension, SBP, DBP, and preeclampsia available in the GWAS Catalog. Genetic variants associated with these traits at p ≤ 1 × 10⁻⁴ were extracted, and variants shared across traits were identified for downstream analyses. For all prioritized variants, we evaluated their potential regulatory effects on nearby genes within a 1 Mb window (cis region) using eQTL analyses.

**Figure 1:**
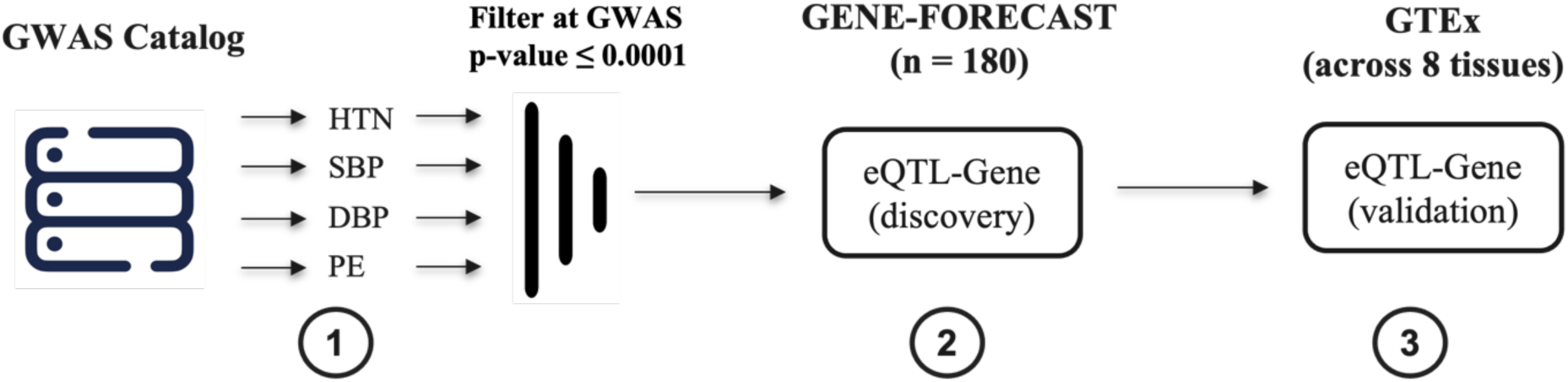
Summary statistics from GWAS of hypertension (HTN), systolic blood pressure (SBP), diastolic blood pressure (DBP), and preeclampsia (PE) were first retrieved from the GWAS Catalog (Step 1). Variants shared across these traits were identified and included in cis-eQTL analysis conducted in whole blood using RNA-seq data from 180 women in the GENE-FORECAST cohort to identify variant-gene associations (Step 2). Finally, identified eQTL-gene pairs were evaluated for replication across eight biologically relevant tissues in GTEx to assess tissue-specific regulatory effects (Step 3).

Cis-eQTL analyses were conducted in whole blood using RNA-seq data of 180 women from the GENE-FORECAST cohort to determine whether shared GWAS variants influence gene expression. This approach provides insight into the regulatory mechanisms through which genetic loci may contribute to both blood pressure regulation and preeclampsia pathophysiology.

Analyses were performed using the R package *MatrixEQTL*, fitting models with normalized mRNA expression as the dependent variable and genotype dosages as the independent variable. Models were adjusted for age, and the top six principal components to account for genetic-ancestry admixture. Variant-gene associations were deemed statistically significant if they reached a false discovery rate (FDR)-adjusted p-value ≤ 0.05. Analyses were restricted to biallelic variants (e.g., single-nucleotide polymorphisms), based on their higher genotyping accuracy and reliability relative to multiallelic variants. This increased precision enhances the robustness of our eQTL findings. Furthermore, the binary allele composition of biallelic variants facilitates interpretation of genetic effects and streamlines identification and characterization of associations between specific alleles and mRNA levels.

Identified eQTL-gene pairs were subsequently evaluated in GTEx across selected biologically relevant tissues, including *Artery Aorta*, *Artery Tibial*, *Artery Coronary*, *Kidney Cortex*, *Whole Blood*, *Uterus*, *Adipose Visceral Omentum*, and *Adrenal Gland*. These tissues were selected based on their established relevance to blood pressure regulation and the pathophysiology of preeclampsia, with an emphasis on vascular, renal, immune, metabolic, and endocrine systems. Arterial tissues capture mechanisms related to vascular tone and remodeling, while kidney cortex reflects renal regulation of blood pressure. Whole blood represents systemic immune and inflammatory processes. Uterine tissue accounts for maternal physiological adaptations during pregnancy, and visceral adipose (omentum) reflects its central role in cardiometabolic regulation and inflammation. Finally, adrenal gland captures endocrine control of blood pressure via steroid hormone pathways. Together, these tissues provide a biologically informed framework to interrogate the functional consequences of shared genetic variants across hypertension and preeclampsia.

Replication in GTEx was defined as a variant–gene association demonstrating (i) a concordant direction of effect (beta) with the discovery analysis, (ii) an FDR-adjusted p-value ≤ 0.05 in the discovery data, and (iii) a nominal p-value ≤ 0.05 in any tissue in GTEx.

### Differential Expression Analysis

To evaluate the downstream phenotypic relevance of genes identified through eQTL mapping, we used the same set of 180 women to test whether expression levels of eQTL-target genes were associated with blood pressure traits, including hypertension status, SBP, and DBP. This step was designed to functionally contextualize genetic regulatory signals by linking genetically influenced gene expression to clinically relevant blood pressure phenotypes, thereby strengthening biological interpretation beyond variant-level associations.

Differential expression analyses were performed using the R library *edgeR*, which models count-based RNA-seq data using negative binomial distributions^14^. For each gene, we fitted models incorporating the trait of interest (hypertension, SBP, or DBP) as the primary predictor, while adjusting for age as a covariate. Tagwise dispersion estimates were used to account for gene-specific variability in expression, improving model fit and inference accuracy. For continuous traits (SBP and DBP), the predictor was included as a continuous variable, whereas hypertension was modeled as a binary variable. Statistical significance was defined using a nominal p-value ≤ 0.05. This analysis enabled the identification of genes whose expression levels are associated with blood pressure phenotypes, providing an additional layer of evidence supporting their potential role in the molecular mechanisms underlying hypertension and related traits.

## RESULTS

### GWAS data

Using the GWAS Catalog trait entries for preeclampsia (EFO 0000668), essential hypertension (EFO 0000537), SBP (EFO 0006335), and DBP (EFO 0006336), we identified and retrieved large-scale GWAS summary statistics for each phenotype. Studies with publicly available summary statistics and adequate genomic coverage were prioritized and harmonized to the GRCh38 reference build to ensure consistency in genomic coordinates across analyses. Genome-wide association study (GWAS) signals were then filtered at a significance threshold of p ≤ 1 × 10⁻⁴. In total, 117 hypertension studies yielded 663,197 variants meeting this threshold, while 112 studies of systolic blood pressure (SBP) identified 1,065,509 variants. For diastolic blood pressure (DBP), 83 studies contributed 841,752 variants, and 20 preeclampsia studies yielded 51,750 variants passing the same criterion. Variants present across all four traits (Preeclampsia, Hypertension, SBP, and DBP) were subsequently retained for downstream cross-trait analyses.

### Shared genetic variants between the four phenotypes

Following extraction of variants meeting the significance threshold (p ≤ 1 × 10⁻⁴) for each phenotype, we performed a cross-trait intersection analysis to identify shared genetic signals across Preeclampsia, Hypertension, SBP, and DBP. This approach yielded 4,792 overlapping variants present across all four phenotypes, representing a core set of shared genetic loci for downstream functional analyses. The overlap structure across traits is illustrated graphically in **Figure 2**. Of the 4,792 shared variants, 4,663 were present in the GENE-FORECAST dataset and had a minor allele frequency (MAF) ≥ 0.01; these variants were retained for downstream eQTL discovery analyses.

**Figure 2:**
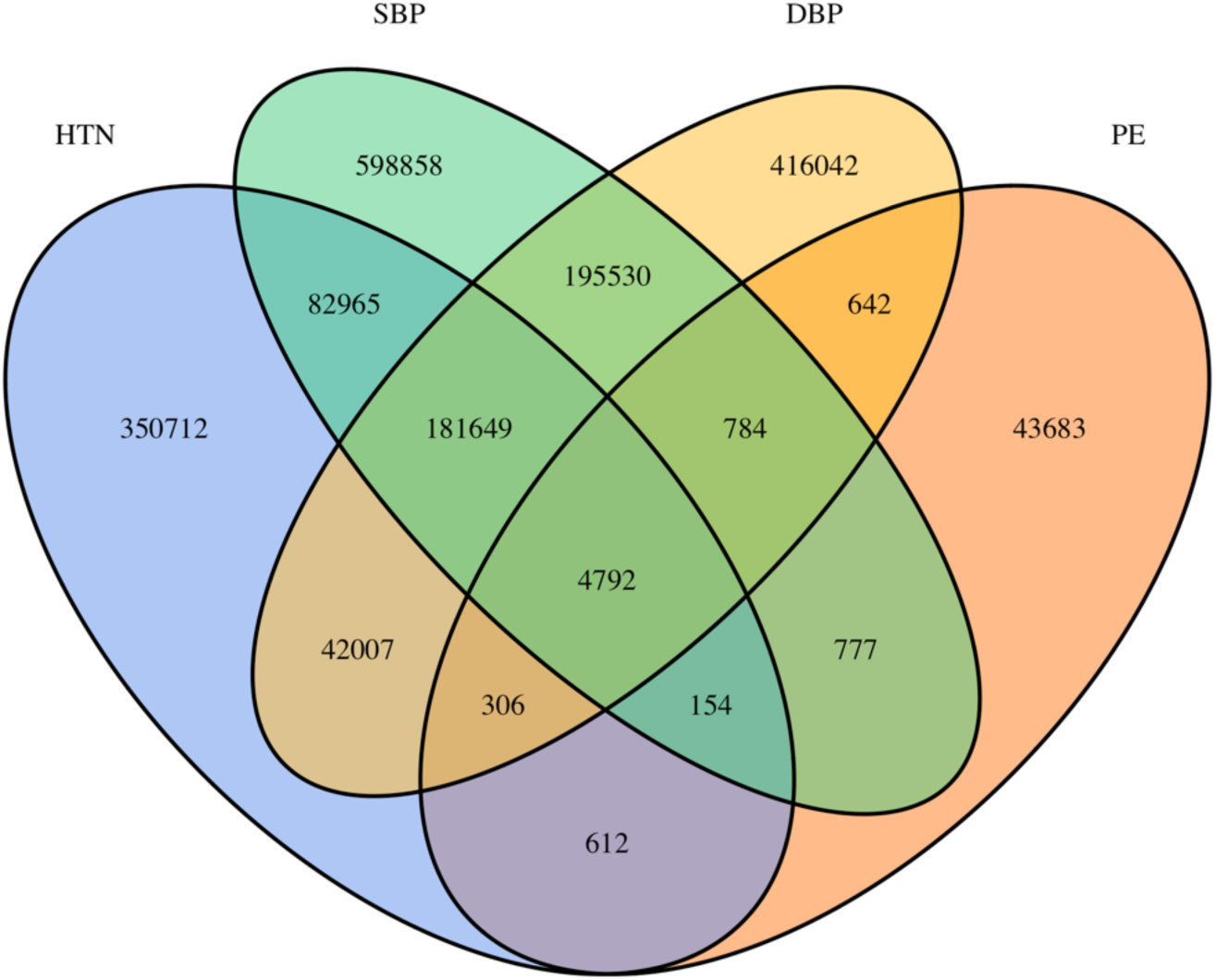
Overlap of variants associated with hypertension (HTN), SBP, DBP, and preeclampsia (PE). Variants meeting the significance threshold (p ≤ 1 × 10⁻⁴) for each phenotype were intersected to identify shared and trait-specific signals. The number of variants in each unique and overlapping region is shown.

### Demographic and clinical characteristics of study participants

We used whole-blood transcriptome and genotype data from 180 women for the eQTL analysis. **Table 1** summarizes participants’ demographic and clinical characteristics, including age, blood pressure, kidney function (estimated glomerular filtration rate, eGFR), anthropometric measures, lipid profile, and behaviors such as smoking and alcohol consumption.

**Table 1:** Demographic, clinical, and socio-behavioral characteristics of the study population (n = 180). Continuous variables are presented as mean ± standard deviation (SD), and categorical variables as counts and proportions.

| Characteristics | Mean/Count | SD/Proportion |
| --- | --- | --- |
| Age (years) | 48 | 12 |
| **Hypertension** |  |  |
| No | 22 | 12% |
| Yes | 158 | 88% |
| Systolic Blood Pressure (mm Hg) | 174 | 37 |
| Diastolic Blood Pressure (mm Hg) | 74 | 10 |
| Estimated Glomerular Filtration Rate (mL/min/1.73 m<sup>2</sup>) | 97 | 21 |
| Body Mass Index | 32 | 7 |
| Low Density Lipoprotein (mg/dL) | 114 | 44 |
| High Density Lipoprotein (mg/dL) | 62 | 14 |
| Triglycerides (mg/dL) | 76 | 38 |
| Glucose (mg/dL) | 102 | 31 |
| C-Reactive Protein (mg/L) | 5 | 8 |
| **Education Level** |  |  |
| ≤ High School | 55 | 31% |
| High School + Vocational Educational | 54 | 30% |
| ≥ Graduate Degree | 71 | 39% |
| **Current Smoker** |  |  |
| No | 170 | 94% |
| Yes | 10 | 6% |

| **Drinks Alcohol** |  |  |
| --- | --- | --- |
| No | 67 | 37% |
| Yes | 113 | 63% |

The study population comprised 180 individuals with a mean age of 48 ± 12 years. Most participants were hypertensive (88%), with only 12% classified as non-hypertensive. Mean systolic and diastolic blood pressures were 174 ± 37 mm Hg and 74 ± 10 mm Hg, respectively. Kidney function was within the normal range, with a mean estimated glomerular filtration rate of 97 ± 21 mL/min/1.73 m². Participants were, on average, in the obese range (BMI: 32 ± 7).

Lipid, metabolic, and inflammatory markers showed variability across the cohort. Socio-behavioral characteristics indicated a diverse educational background, with the majority of participants being non-smokers and approximately two-thirds reporting alcohol consumption.

### eQTL results and validation in GTEx

The eQTL analysis was performed on expression profiles of 17,947 protein-coding genes, testing associations with 4,663 GWAS variants shared among Preeclampsia, Hypertension, SBP, and DBP that were present in our whole-genome sequencing data with a MAF ≥ 0.05. Using an FDR-adjusted p-value ≤ 0.05 criterion, we identified 1,837 eQTL-mRNA associations, involving 1,070 unique variants and 78 unique protein-coding genes. The full list of eQTL-Gene pairs is reported in **Table S1** in the **Supplemental Material**.

To place the identified eQTL-gene associations in a broader biological context, we evaluated their presence in GTEx. GTEx represents one of the most extensive resources for characterizing cis-eQTLs across human tissues. Notably, whole blood, the tissue used in our discovery analysis, is among the most well-powered tissues within the GTEx dataset, enabling robust comparison and validation of regulatory signals.

For replication analyses in GTEx, we prioritized tissues with established relevance to blood pressure regulation and preeclampsia pathophysiology. These included arterial tissues (aorta, tibial, and coronary) to capture vascular tone and remodeling, and kidney cortex to reflect renal control of blood pressure. Whole blood was included to replicate regulatory signals from the primary discovery tissue and to capture systemic immune and inflammatory processes. Uterus was included to represent maternal physiological adaptations during pregnancy, and visceral adipose (omentum) to capture its role in cardiometabolic regulation and inflammation. Finally, adrenal gland was included to account for endocrine regulation of blood pressure via steroid hormone pathways.

Replication analyses in GTEx confirmed 645 eQTL-gene associations involving 569 variants and 24 genes across the eight selected tissues (**Table S2** in the **Supplemental Material)**. Notably, several signals, including 19 of the 24 loci, showed significant effects across multiple vascular, metabolic, and endocrine tissues, supporting the robustness and cross-tissue relevance of these regulatory associations (**Table 2**).

**Table 2:** Genes for which eQTL-gene pairs were identified, along with the number and types of tissues in which these regulatory associations were observed.

| **Gene** | **Number of tissues where eQTL-Gene pair is observed** | **Tissue** |
| --- | --- | --- |
| H3C6 | 8 | Adipose Visceral Omentum; Adrenal Gland; Artery Aorta; Artery Coronary; Artery Tibial; Kidney Cortex; Uterus; Whole Blood |
| HLA-DQA2 | 8 | Adipose Visceral Omentum; Adrenal Gland; Artery Aorta; Artery Coronary; Artery Tibial; Kidney Cortex; Uterus; Whole Blood |
| HLA-DRB1 | 8 | Adipose Visceral Omentum; Adrenal Gland; Artery Aorta; Artery Coronary; Artery Tibial; Kidney Cortex; Uterus; Whole Blood |
| LRRC37A | 8 | Adipose Visceral Omentum; Adrenal Gland; Artery Aorta; Artery Coronary; Artery Tibial; Kidney Cortex; Uterus; Whole Blood |
| RPS26 | 8 | Adipose Visceral Omentum; Adrenal Gland; Artery Aorta; Artery Coronary; Artery Tibial; Kidney Cortex; Uterus; Whole Blood |
| HLA-C | 7 | Adipose Visceral Omentum; Adrenal Gland; Artery Aorta; Artery Coronary; Artery Tibial; Uterus; Whole Blood |
| HLA-DQB1 | 7 | Adipose Visceral Omentum; Adrenal Gland; Artery Aorta; Artery Coronary; Artery Tibial; Uterus; Whole Blood |
| HLA-DRB5 | 7 | Adipose Visceral Omentum; Adrenal Gland; Artery Aorta; Artery Coronary; Artery Tibial; Uterus; Whole Blood |
| MICA | 7 | Adipose Visceral Omentum; Adrenal Gland; Artery Aorta; Artery Coronary; Artery Tibial; Kidney Cortex; Whole Blood |
| HLA-DQA1 | 6 | Adipose Visceral Omentum; Artery Aorta; Artery Coronary; Artery Tibial; Kidney Cortex; Whole Blood |
| TMEM116 | 6 | Adipose Visceral Omentum; Adrenal Gland; Artery Aorta; Artery Coronary; Artery Tibial; Whole Blood |
| TUFM | 6 | Adipose Visceral Omentum; Artery Aorta; Artery Coronary; Artery Tibial; Uterus; Whole Blood |
| FADS2 | 5 | Adipose Visceral Omentum; Artery Aorta; Artery Coronary; Artery Tibial; Whole Blood |
| SULT1A1 | 5 | Adipose Visceral Omentum; Artery Aorta; Artery Coronary; Artery Tibial; Whole Blood |
| HLA-B | 4 | Adipose Visceral Omentum; Artery Aorta; Artery Coronary; Artery Tibial |
| MAPKAPK5 | 4 | Adipose Visceral Omentum; Artery Aorta; Artery Coronary; Artery Tibial |
| PSORS1C1 | 4 | Adipose Visceral Omentum; Artery Aorta; Artery Coronary; Artery Tibial |
| SGF29 | 4 | Adipose Visceral Omentum; Artery Aorta; Artery Tibial; Whole Blood |
| CCHCR1 | 3 | Adipose Visceral Omentum; Artery Aorta; Whole Blood |
| C4B | 1 | Whole Blood |
| FADS1 | 1 | Whole Blood |
| NOTCH4 | 1 | Whole Blood |
| SULT1A2 | 1 | Whole Blood |
| ZSWIM5 | 1 | Artery Tibial |

Across tissues, eQTL-gene associations showed clear enrichment in vascular and metabolically active tissues (**Table 3**). Artery Tibial (n = 609), Adipose Visceral Omentum (n = 595), and Whole Blood (n = 578) had the most associations, followed by Artery Aorta and Artery Coronary, indicating a strong vascular regulatory component. In contrast, Kidney Cortex and Uterus had fewer associations, suggesting more tissue-restricted regulatory effects.

**Table 3:** The number of significant eQTL-gene pairs identified in each tissue, ranked by total associations, along with the corresponding genes involved in these regulatory relationships.

| Tissue | Number of eQTL-Gene pairs | Genes |
| --- | --- | --- |
| Artery<br>Tibial | 609 | HLA-B; FADS2; H3C6; HLA-DQA2; RPS26; MICA; LRRC37A;<br>HLA-DQB1; SGF29; HLA-DQA1; HLA-DRB1; TMEM116;<br>HLA-C; SULT1A1; HLA-DRB5; TUFM; PSORS1C1;<br>MAPKAPK5; ZSWIM5 |
| Adipose<br>Visceral<br>Omentum | 595 | HLA-B; FADS2; H3C6; HLA-DQA2; RPS26; MICA; LRRC37A;<br>HLA-DQB1; SGF29; HLA-DQA1; HLA-DRB1; TMEM116;<br>HLA-C; CCHCR1; SULT1A1; HLA-DRB5; TUFM; PSORS1C1;<br>MAPKAPK5 |
| Whole<br>Blood | 578 | NOTCH4; FADS2; FADS1; H3C6; HLA-DQA2; RPS26; MICA;<br>LRRC37A; HLA-DQB1; SGF29; SULT1A2; HLA-DQA1;<br>HLA-DRB1; TMEM116; HLA-C; CCHCR1; C4B; SULT1A1;<br>HLA-DRB5; TUFM |
| Artery<br>Aorta | 552 | HLA-B; FADS2; H3C6; HLA-DQA2; RPS26; MICA; LRRC37A;<br>HLA-DQB1; SGF29; HLA-DQA1; HLA-DRB1; TMEM116;<br>HLA-C; CCHCR1; SULT1A1; HLA-DRB5; TUFM; PSORS1C1;<br>MAPKAPK5 |
| Artery<br>Coronary | 543 | HLA-B; FADS2; H3C6; HLA-DQA2; RPS26; MICA; LRRC37A;<br>HLA-DQB1; HLA-DQA1; HLA-DRB1; TMEM116; HLA-C;<br>SULT1A1; HLA-DRB5; TUFM; PSORS1C1; MAPKAPK5 |
| Adrenal<br>Gland | 466 | H3C6; HLA-DQA2; RPS26; MICA; LRRC37A; HLA-DQB1;<br>HLA-DRB1; TMEM116; HLA-C; HLA-DRB5 |
| Uterus | 131 | H3C6; HLA-DQA2; RPS26; LRRC37A; HLA-DQB1;<br>HLA-DRB1; HLA-C; HLA-DRB5; TUFM |
| Kidney<br>Cortex | 84 | H3C6; HLA-DQA2; RPS26; MICA; LRRC37A; HLA-DQA1;<br>HLA-DRB1 |

Notably, several genes, particularly within the HLA region (e.g., HLA-DQA2, HLA-DRB1, HLA-DQB1), as well as RPS26, LRRC37A, and MICA, were consistently observed across multiple tissues, highlighting shared immune and inflammatory regulatory mechanisms. Additional tissue-specific signals (e.g., NOTCH4 and FADS1 in whole blood, ZSWIM5 in tibial artery) suggest complementary, context-dependent regulatory effects.

### Association of eQTL-target gene expression with blood pressure traits

To assess the functional relevance of genes identified through eQTL mapping, we evaluated the association between gene expression levels and blood pressure traits, including hypertension, SBP, and DBP, in the same set of 180 women. All 24 genes with at least one associated eQTL were tested. This analysis was designed to link genetically regulated gene expression to clinically measurable phenotypes, thereby strengthening the interpretation of the eQTL findings.

Among the 24 genes examined, 3 demonstrated statistically significant associations (**Table S3** in the **Supplemental Material)**. For hypertension, Complement Component 4B (C4B) (logFC = 0.94, p = 0.004) and Major Histocompatibility Complex, Class I, C (HLA-C) (logFC = -0.90, p = 0.026) were significantly associated, with opposing directions of effect. For SBP, both Complement Component 4B (C4B) (logFC = 0.0062, p = 0.023) and Major Histocompatibility Complex, Class I, C (HLA-C) (logFC = -0.0070, p = 0.047) remained significant, showing consistent directionality with hypertension. For DBP, Major Histocompatibility Complex, Class I, C (HLA-C) (logFC = -0.0405, p = 0.001) and Major Histocompatibility Complex, Class II, DQ Beta 1 (HLA-DQB1) (logFC = 0.0260, p = 0.01) were significantly associated, indicating a trait-specific effect for HLA-DQB1.

For the two continuous traits (SBP and DBP), the reported logFC values represent the change in gene expression associated with a one-unit increase in blood pressure (1 mmHg). Therefore, although the effect sizes appear numerically small, they reflect incremental changes per mmHg and should be interpreted cumulatively over clinically meaningful differences in blood pressure.

The remaining 21 genes were not significantly associated with any of the traits examined. These results suggest that only a subset of eQTL-target genes shows detectable expression-phenotype relationships in this dataset, highlighting a limited number of candidates with convergent genetic and functional evidence.

### Integration of shared genetic signals with gene expression and blood pressure traits

To facilitate interpretation of the results, in **Table 4**, we summarized the combined evidence linking GWAS associations, cis-eQTL effects, and gene expression-trait relationships for the three genes: C4B, HLA-C, and HLA-DQB1.

**Table 4:** This table synthesizes the direction and consistency of genetic associations (GWAS) and regulatory effects (cis-eQTLs) for C4B, HLA-C, and HLA-DQB1. For each gene, the number of associated eQTL variants and their effects on gene expression are reported, along with the number and direction of GWAS associations for hypertension, preeclampsia, systolic blood pressure, and diastolic blood pressure. Arrows indicate the direction of effect (↑ increase, ↓ decrease). Variant counts should be interpreted as representing locus-level signals rather than independent associations.

| Gene symbol | C4B | HLA-C | HLA-DQB1 |
| --- | --- | --- | --- |
| # eQTL variants | 4 | 3 | 32 |
| eQTL effect gene | Up-regulation (4 eQTL ↑) | Mixed (2 eQTL ↑ 1 eQTL ↓) | Down-regulation (32 eQTL ↓) |
| GWAS effect on HTN | Risk-increasing (12 studies ↑) | Risk-increasing (3 studies ↑) | Risk-increasing (194 studies ↑) |
| GWAS effect on PE | Risk-increasing (4 studies ↑) | Risk-increasing (4 studies ↑) | Risk-increasing (33 studies ↑) |
| GWAS effect on SBP | Increasing level (24 studies ↑) | Increasing level (19 studies ↑) | Increasing level (406 studies ↑) |
| GWAS effect on DBP | Increasing level (4 studies ↑) | Increasing level (69 studies ↑) | Increasing level (803 studies ↑) |

Across all three genes, GWAS findings consistently indicated risk-increasing effects for hypertension and preeclampsia, as well as increasing effects on SBP and DBP levels. These associations are represented by multiple variants; however, many of these variants are in linkage disequilibrium, particularly within the HLA region. Accordingly, the number of associated variants or studies should be interpreted as reflecting underlying locus-level signals captured by correlated variants, rather than independent associations. This is most evident for HLA-DQB1, where the large number of associated variants likely reflects a limited number of LD-driven signals within a highly structured genomic region.

The eQTL results further highlight gene-specific regulatory patterns. For C4B, all identified eQTL variants showed consistent upregulation of gene expression, suggesting a coherent regulatory effect at this locus. In contrast, HLA-DQB1 showed a uniform downregulatory profile, with all associated eQTL variants linked to decreased expression, consistent with a shared underlying regulatory signal. HLA-C exhibited a mixed pattern of eQTL effects, indicating either multiple signals or more complex regulatory mechanisms.

When considered together, the directionality of GWAS and eQTL effects supports locus-level convergence between genetic risk and gene regulation. For C4B, risk-increasing GWAS effects align with increased expression, whereas for HLA-DQB1, risk-increasing effects coincide with decreased expression. In contrast, the mixed regulatory profile observed for HLA-C, despite consistent GWAS directionality, suggests a more complex relationship between genetic variation and gene expression at this locus.

All 39 eQTLs associated with C4B, HLA-C, and HLA-DQB1 were common variants, with minor allele frequencies (MAF) ranging from 0.12 to 0.49. Collectively, these variants were linked to multiple traits, including hypertension (209 associations), preeclampsia (41 associations), systolic blood pressure (449 associations), and diastolic blood pressure (876 associations). Overall, 993 of the 1,575 reported associations (63%) reached genome-wide significance (p ≤ 5.0 × 10⁻⁸). Full details of all variant-gene-trait relationships are provided in **Table S4** in the **Supplemental Material**.

## DISCUSSION

### Main findings

In this study, we integrated genome-wide association signals for hypertension, preeclampsia, and systolic and diastolic blood pressure with transcriptomic data to identify shared regulatory mechanisms linking genetic variation to clinically relevant phenotypes. Among 4,792 shared variants across the four traits, we identified 1,837 eQTL–gene associations in whole blood, of which 645 associations involving 24 genes were replicated across multiple tissues in the GTEx project.

Among these, only three genes (C4B, HLA-C, and HLA-DQB1) demonstrated consistent evidence across all three analytical layers: (i) GWAS associations with hypertension, preeclampsia, and systolic and diastolic blood pressure; (ii) cis-eQTL effects on gene expression; and (iii) significant associations between gene expression and blood pressure traits in the same individuals. These genes, therefore, represent a subset of loci where genetic variation, gene regulation, and phenotype converge, providing stronger functional support than variant-level associations alone.

### From shared genetic architecture to functional mechanisms

A key contribution of this study is extending GWAS findings beyond association toward mechanistic interpretation. While GWAS have identified hundreds of loci associated with blood pressure traits and, more recently, preeclampsia^6, 8^, the functional consequences of most variants remain unclear, particularly given their predominant localization in non-coding regions^10^.

By integrating GWAS signals with eQTL mapping and downstream expression-trait analyses, we demonstrate that only a limited number of loci exhibit coherent effects across all levels, supporting a model in which shared genetic architecture between hypertension and preeclampsia is driven by a restricted set of functionally active loci rather than a diffuse polygenic background.

Notably, the associations observed at each locus are represented by sets of variants in linkage disequilibrium, particularly within the HLA region. Therefore, our findings should be interpreted at the locus level, reflecting underlying regulatory signals captured by correlated variants rather than by independent effects.

### Gene-specific insights into convergent genetic and regulatory mechanisms

C4B emerged as the most coherent signal across all analytical layers. GWAS associations consistently indicated risk-increasing effects, and eQTL analyses showed uniform upregulation of gene expression. Expression levels were positively associated with hypertension and SBP. This directional concordance supports a model in which increased C4B expression is associated with elevated blood pressure and an increased risk of hypertension.

C4B encodes a component of the complement system, which plays a central role in innate immunity and inflammation^15^. Dysregulation of complement activation has been implicated in vascular dysfunction and hypertensive disorders, including preeclampsia, as a key driver of inflammation and endothelial damage^16^. Excessive activation or insufficient regulation of complement components triggers inflammation, disrupts angiogenesis, and results in placental insufficiency and maternal vascular dysfunction^17, 18^.

Our findings position C4B as a strong candidate linking genetic variation to immune-mediated vascular mechanisms underlying both hypertension and preeclampsia.

HLA-C demonstrated a more complex regulatory architecture. While GWAS associations consistently indicated increased risk across traits, eQTL effects were mixed in direction, suggesting heterogeneous regulatory influences or multiple underlying signals. Despite this, gene expression was consistently negatively associated with hypertension, SBP, and DBP.

This apparent discordance highlights an important point: consistent phenotype associations can arise even when regulatory effects are heterogeneous, particularly in genomic regions characterized by complex LD and multiple regulatory elements. The HLA region is known for its extensive polymorphism and regulatory complexity^19^, which complicates the interpretation of variant-level effects.

HLA-C is a key component of antigen presentation and immune regulation. Variation in HLA-C has been implicated in immune-mediated diseases and in maternal-fetal immune interactions relevant to preeclampsia^20, 21^. Reduced expression of HLA-C may influence immune tolerance and inflammatory responses, potentially contributing to vascular dysfunction^22, 23^.

Thus, HLA-C represents a locus where genetic and phenotypic signals converge despite complex underlying regulation, reinforcing its relevance to shared disease mechanisms.

HLA-DQB1 showed the strongest GWAS signal, with a large number of associated variants across all traits. However, this reflects the extended LD structure of the HLA region rather than a high number of independent associations. At the regulatory level, eQTL effects were uniformly downregulatory, indicating a consistent underlying signal.

In contrast to C4B and HLA-C, HLA-DQB1 expression was specifically associated with DBP, but not with hypertension or SBP, suggesting a trait-specific downstream effect. Consistent with this observation, the majority of GWAS associations linked to variants influencing HLA-DQB1 expression were also related to DBP, accounting for approximately 60% of all GWAS associations across the four traits, further supporting a preferential role of this locus in diastolic blood pressure regulation. This indicates that even when genetic and regulatory signals are strong, their phenotypic impact may be selective.

HLA-DQB1 is involved in antigen presentation and adaptive immune responses and has been implicated in multiple autoimmune and inflammatory conditions^24^. Immune dysregulation has been increasingly recognized in the pathophysiology of both hypertension and preeclampsia^25, 26^.

Our findings suggest that HLA-DQB1 contributes to blood pressure regulation through specific pathways that influence diastolic blood pressure, highlighting the importance of distinguishing between related yet distinct phenotypes.

### Biological convergence on immune and inflammatory pathways

A notable finding of this study is that all three prioritized genes, C4B, HLA-C, and HLA-DQB1, are involved in immune function. This convergence supports the hypothesis that immune and inflammatory mechanisms constitute a shared biological axis linking hypertension and preeclampsia.

This is consistent with prior studies demonstrating the role of immune activation, complement pathways, and inflammatory signaling in vascular dysfunction and blood pressure regulation ^18, 27–29^. In preeclampsia, immune maladaptation and systemic inflammation are central features, while in hypertension, chronic low-grade inflammation contributes to endothelial dysfunction and vascular remodeling^25, 30, 31^.

Thus, our integrative analysis provides genetic and transcriptomic evidence supporting immune-mediated mechanisms as a common pathway underlying both conditions

### Cross-tissue validation supports systemic regulatory effects

Although our primary eQTL analysis was conducted in whole blood, we validated findings across multiple tissues using GTEx, including vascular (arterial), renal, metabolic, endocrine, and reproductive tissues. A substantial proportion of eQTL-gene associations replicated across these tissues, with several genes, including HLA-C and HLA-DQB1, demonstrating broad cross-tissue regulatory activity.

This cross-tissue consistency indicates that the identified regulatory signals are not limited to a single tissue context but instead reflect systemic regulatory mechanisms relevant to blood pressure physiology. The enrichment of signals in arterial and metabolically active tissues further supports the biological plausibility of these findings.

From a translational perspective, the convergence of findings on complement and HLA-mediated pathways raises the possibility that immune-modulating therapies currently under investigation for autoimmune conditions could be repurposed for hypertensive disorders. Complement inhibitors such as eculizumab have already been explored in severe preeclampsia on a compassionate-use basis, with one case demonstrating marked clinical improvement and prolongation of pregnancy by 17 days^32^, and our findings provide a genetic and transcriptomic rationale for further investigation of this therapeutic axis^16^. More broadly, immune modulation has been proposed as a complementary strategy for difficult-to-treat hypertension, given the established role of innate and adaptive immune responses in driving vascular and end-organ damage^28, 33^. Additionally, the identification of common variants with regulatory effects across multiple tissues suggests that peripheral blood-based biomarkers, particularly C4B and HLA-C expression levels, could be explored as predictive markers linking hypertension risk to future preeclampsia susceptibility. Notably, a recent systematic review of proteomics-based biomarkers reported that serum complement C4b levels were consistently reduced in early gestation among women who later developed preeclampsia, supporting C4B as a candidate for early detection^34^.

### Strengths and limitations

This study has several strengths. First, the integration of genotype, gene expression, and phenotype data within the same individuals minimizes confounding and strengthens interpretation. Second, the use of multi-tissue validation in GTEx enhances the robustness and generalizability of the findings. Third, the focus on shared variants across hypertension, preeclampsia, and systolic and diastolic blood pressure provides a novel perspective on common disease mechanisms.

Limitations should also be considered. The cross-sectional design precludes causal inference regarding the relationship between gene expression and traits. Only 3 of the 24 genes tested showed significant associations between gene expression and blood pressure traits. Most regulatory signals identified through eQTL analyses do not translate into detectable phenotypic effects, at least within the sample size and tissue context of this study. This observation indicates the need for multi-layered validation frameworks, as reliance on GWAS or eQTL data alone may overestimate the number of functionally relevant loci. Finally, the relationship between gene expression and preeclampsia could not be evaluated, as preeclampsia status was not assessed among the study participants.

## PERSPECTIVES

Our findings demonstrate that integrating GWAS signals across hypertension, blood pressure traits, and preeclampsia with transcriptomic data can identify a limited number of loci where genetic variation, gene regulation, and clinical phenotypes converge. The emergence of C4B, HLA-C, and HLA-DQB1 as the only genes exhibiting this full mechanistic coherence points to immune and inflammatory pathways as a central biological axis shared between chronic and pregnancy-related hypertensive disorders. This has several broader implications. First, it suggests that therapeutic strategies targeting complement activation or HLA-mediated immune responses may have relevance beyond single-disease contexts, potentially benefiting both hypertension management and preeclampsia prevention. Second, the trait-specific association of HLA-DQB1 with diastolic blood pressure underscores the importance of disaggregating blood pressure components in genetic studies, as shared upstream regulation does not necessarily produce uniform downstream effects. Third, our multi-layered analytical framework, requiring convergence across GWAS, eQTL, and expression-trait analyses, provides a generalizable approach for prioritizing functionally relevant loci among the large number of statistical associations generated by GWAS.

## NOVELTY AND RELEVANCE

### What Is New?

- This study identifies C4B, HLA-C, and HLA-DQB1 as the only loci demonstrating full mechanistic coherence, from genetic variant to gene regulation to blood pressure phenotype, across hypertension, systolic and diastolic blood pressure, and preeclampsia.
- HLA-DQB1 expression was specifically associated with diastolic blood pressure, revealing trait-specific downstream effects despite shared upstream genetic signals.

### What Is Relevant?

- These findings implicate immune regulatory mechanisms, particularly complement activation and antigen presentation, as shared biological pathways linking chronic hypertension and preeclampsia.
- The multi-layered analytical framework used here offers a strategy for moving beyond GWAS associations toward functionally interpretable targets.

## SOURCES OF FUNDING

This work was supported by the Chan Zuckerberg Initiative’s Foundation to Accelerate Precision Health Program and Advance Genomics Research at Meharry Medical College (CZIF2022-007043) and by the NIMHD grant U54MD007593 to build research capacity at Meharry Medical College.

## DISCLOSURES

None.

## DECLARATION OF GENERATIVE AI AND AI-ASSISTED TECHNOLOGIES IN THE WRITING PROCESS

During the preparation of this work, the author(s) used Grammarly in order to check and correct language spelling and grammar. After using this tool/service, the author(s) reviewed and edited the content as needed and take(s) full responsibility for the content of the publication.

## SUPPLEMENTAL MATERIAL

Supplemental Tables S1–S4

## Data Availability

Data will not be shared or will be available upon request.

## Notes

### Competing Interest Statement

The authors have declared no competing interest.

### Funding Statement

This work was supported by the Chan Zuckerberg Initiatives Foundation to Accelerate Precision Health Program and Advance Genomics Research at Meharry Medical College (CZIF2022-007043) and by the NIMHD grant U54MD007593 to build research capacity at Meharry Medical College.

### Author Declarations

The study protocol was approved by the Institutional Review Boards of the National Institutes of Health and Meharry Medical College. Written informed consent was obtained from all participants prior to enrollment, and all study procedures adhered to institutional and regulatory requirements.

### Summary of Updates

First author's name was corrected because it was missing the middle name.

## REFERENCES

1. World Health Organization. Hypertension Geneva: World Health Organization; 2025 [cited 2026 2026 Mar 24]. Available from: https://www.who.int/news-room/fact-sheets/detail/hypertension.

2. Mills KT, Stefanescu A, He J. The global epidemiology of hypertension. Nat Rev Nephrol. 2020;16(4):223–37. Epub 20200205. doi: 10.1038/s41581-019-0244-2. PubMed PMID: 32024986; PMCID: PMC7998524.

3. Karadzov Orlic N, Joksić I. Preeclampsia pathogenesis and prediction - where are we now: the focus on the role of galectins and miRNAs. Hypertens Pregnancy. 2025;44(1):2470626. Epub 20250227. doi: 10.1080/10641955.2025.2470626. PubMed PMID: 40012493.

4. Maas A, Rosano G, Cifkova R, Chieffo A, van Dijken D, Hamoda H, Kunadian V, Laan E, Lambrinoudaki I, Maclaran K, Panay N, Stevenson JC, van Trotsenburg M, Collins P. Cardiovascular health after menopause transition, pregnancy disorders, and other gynaecologic conditions: a consensus document from European cardiologists, gynaecologists, and endocrinologists. Eur Heart J. 2021;42(10):967–84. doi: 10.1093/eurheartj/ehaa1044. PubMed PMID: 33495787; PMCID: PMC7947184.

5. Verma A, Huffman JE, Rodriguez A, Conery M, Liu M, Ho YL, Kim Y, Heise DA, Guare L, Panickan VA, Garcon H, Linares F, Costa L, Goethert I, Tipton R, Honerlaw J, Davies L, Whitbourne S, Cohen J, Posner DC, Sangar R, Murray M, Wang X, Dochtermann DR, Devineni P, Shi Y, Nandi TN, Assimes TL, Brunette CA, Carroll RJ, Clifford R, Duvall S, Gelernter J, Hung A, Iyengar SK, Joseph J, Kember R, Kranzler H, Kripke CM, Levey D, Luoh SW, Merritt VC, Overstreet C, Deak JD, Grant SFA, Polimanti R, Roussos P, Shakt G, Sun YV, Tsao N, Venkatesh S, Voloudakis G, Justice A, Begoli E, Ramoni R, Tourassi G, Pyarajan S, Tsao P, O’Donnell CJ, Muralidhar S, Moser J, Casas JP, Bick AG, Zhou W, Cai T, Voight BF, Cho K, Gaziano JM, Madduri RK, Damrauer S, Liao KP. Diversity and scale: Genetic architecture of 2068 traits in the VA Million Veteran Program. Science. 2024;385(6706):eadj1182. Epub 20240719. doi: 10.1126/science.adj1182. PubMed PMID: 39024449; PMCID: PMC12857194.

6. Tyrmi JS, Kaartokallio T, Lokki AI, Jääskeläinen T, Kortelainen E, Ruotsalainen S, Karjalainen J, Ripatti S, Kivioja A, Laisk T, Kettunen J, Pouta A, Kivinen K, Kajantie E, Heinonen S, Kere J, Laivuori H. Genetic Risk Factors Associated With Preeclampsia and Hypertensive Disorders of Pregnancy. JAMA Cardiol. 2023;8(7):674–83. doi: 10.1001/jamacardio.2023.1312. PubMed PMID: 37285119; PMCID: PMC10248811.

7. Keaton JM, Kamali Z, Xie T, Vaez A, Williams A, Goleva SB, Ani A, Evangelou E, Hellwege JN, Yengo L, Young WJ, Traylor M, Giri A, Zheng Z, Zeng J, Chasman DI, Morris AP, Caulfield MJ, Hwang SJ, Kooner JS, Conen D, Attia JR, Morrison AC, Loos RJF, Kristiansson K, Schmidt R, Hicks AA, Pramstaller PP, Nelson CP, Samani NJ, Risch L, Gyllensten U, Melander O, Riese H, Wilson JF, Campbell H, Rich SS, Psaty BM, Lu Y, Rotter JI, Guo X, Rice KM, Vollenweider P, Sundström J, Langenberg C, Tobin MD, Giedraitis V, Luan J, Tuomilehto J, Kutalik Z, Ripatti S, Salomaa V, Girotto G, Trompet S, Jukema JW, van der Harst P, Ridker PM, Giulianini F, Vitart V, Goel A, Watkins H, Harris SE, Deary IJ, van der Most PJ, Oldehinkel AJ, Keavney BD, Hayward C, Campbell A, Boehnke M, Scott LJ, Boutin T, Mamasoula C, Järvelin MR, Peters A, Gieger C, Lakatta EG, Cucca F, Hui J, Knekt P, Enroth S, De Borst MH, Polašek O, Concas MP, Catamo E, Cocca M, Li-Gao R, Hofer E, Schmidt H, Spedicati B, Waldenberger M, Strachan DP, Laan M, Teumer A, Dörr M, Gudnason V, Cook JP, Ruggiero D, Kolcic I, Boerwinkle E, Traglia M, Lehtimäki T, Raitakari OT, Johnson AD, Newton-Cheh C, Brown MJ, Dominiczak AF, Sever PJ, Poulter N, Chambers JC, Elosua R, Siscovick D, Esko T, Metspalu A, Strawbridge RJ, Laakso M, Hamsten A, Hottenga JJ, de Geus E, Morris AD, Palmer CNA, Nolte IM, Milaneschi Y, Marten J, Wright A, Zeggini E, Howson JMM, O’Donnell CJ, Spector T, Nalls MA, Simonsick EM, Liu Y, van Duijn CM, Butterworth AS, Danesh JN, Menni C, Wareham NJ, Khaw KT, Sun YV, Wilson PWF, Cho K, Visscher PM, Denny JC, Levy D, Edwards TL, Munroe PB, Snieder H, Warren HR. Genome-wide analysis in over 1 million individuals of European ancestry yields improved polygenic risk scores for blood pressure traits. Nat Genet. 2024;56(5):778–91. Epub 20240430. doi: 10.1038/s41588-024-01714-w. PubMed PMID: 38689001; PMCID: PMC11096100.

8. Evangelou E, Warren HR, Mosen-Ansorena D, Mifsud B, Pazoki R, Gao H, Ntritsos G, Dimou N, Cabrera CP, Karaman I, Ng FL, Evangelou M, Witkowska K, Tzanis E, Hellwege JN, Giri A, Velez Edwards DR, Sun YV, Cho K, Gaziano JM, Wilson PWF, Tsao PS, Kovesdy CP, Esko T, Mägi R, Milani L, Almgren P, Boutin T, Debette S, Ding J, Giulianini F, Holliday EG, Jackson AU, Li-Gao R, Lin WY, Luan J, Mangino M, Oldmeadow C, Prins BP, Qian Y, Sargurupremraj M, Shah N, Surendran P, Thériault S, Verweij N, Willems SM, Zhao JH, Amouyel P, Connell J, de Mutsert R, Doney ASF, Farrall M, Menni C, Morris AD, Noordam R, Paré G, Poulter NR, Shields DC, Stanton A, Thom S, Abecasis G, Amin N, Arking DE, Ayers KL, Barbieri CM, Batini C, Bis JC, Blake T, Bochud M, Boehnke M, Boerwinkle E, Boomsma DI, Bottinger EP, Braund PS, Brumat M, Campbell A, Campbell H, Chakravarti A, Chambers JC, Chauhan G, Ciullo M, Cocca M, Collins F, Cordell HJ, Davies G, de Borst MH, de Geus EJ, Deary IJ, Deelen J, Del Greco MF, Demirkale CY, Dörr M, Ehret GB, Elosua R, Enroth S, Erzurumluoglu AM, Ferreira T, Frånberg M, Franco OH, Gandin I, Gasparini P, Giedraitis V, Gieger C, Girotto G, Goel A, Gow AJ, Gudnason V, Guo X, Gyllensten U, Hamsten A, Harris TB, Harris SE, Hartman CA, Havulinna AS, Hicks AA, Hofer E, Hofman A, Hottenga JJ, Huffman JE, Hwang SJ, Ingelsson E, James A, Jansen R, Jarvelin MR, Joehanes R, Johansson Å, Johnson AD, Joshi PK, Jousilahti P, Jukema JW, Jula A, Kähönen M, Kathiresan S, Keavney BD, Khaw KT, Knekt P, Knight J, Kolcic I, Kooner JS, Koskinen S, Kristiansson K, Kutalik Z, Laan M, Larson M, Launer LJ, Lehne B, Lehtimäki T, Liewald DCM, Lin L, Lind L, Lindgren CM, Liu Y, Loos RJF, Lopez LM, Lu Y, Lyytikäinen LP, Mahajan A, Mamasoula C, Marrugat J, Marten J, Milaneschi Y, Morgan A, Morris AP, Morrison AC, Munson PJ, Nalls MA, Nandakumar P, Nelson CP, Niiranen T, Nolte IM, Nutile T, Oldehinkel AJ, Oostra BA, O’Reilly PF, Org E, Padmanabhan S, Palmas W, Palotie A, Pattie A, Penninx B, Perola M, Peters A, Polasek O, Pramstaller PP, Nguyen QT, Raitakari OT, Ren M, Rettig R, Rice K, Ridker PM, Ried JS, Riese H, Ripatti S, Robino A, Rose LM, Rotter JI, Rudan I, Ruggiero D, Saba Y, Sala CF, Salomaa V, Samani NJ, Sarin AP, Schmidt R, Schmidt H, Shrine N, Siscovick D, Smith AV, Snieder H, Sõber S, Sorice R, Starr JM, Stott DJ, Strachan DP, Strawbridge RJ, Sundström J, Swertz MA, Taylor KD, Teumer A, Tobin MD, Tomaszewski M, Toniolo D, Traglia M, Trompet S, Tuomilehto J, Tzourio C, Uitterlinden AG, Vaez A, van der Most PJ, van Duijn CM, Vergnaud AC, Verwoert GC, Vitart V, Völker U, Vollenweider P, Vuckovic D, Watkins H, Wild SH, Willemsen G, Wilson JF, Wright AF, Yao J, Zemunik T, Zhang W, Attia JR, Butterworth AS, Chasman DI, Conen D, Cucca F, Danesh J, Hayward C, Howson JMM, Laakso M, Lakatta EG, Langenberg C, Melander O, Mook-Kanamori DO, Palmer CNA, Risch L, Scott RA, Scott RJ, Sever P, Spector TD, van der Harst P, Wareham NJ, Zeggini E, Levy D, Munroe PB, Newton-Cheh C, Brown MJ, Metspalu A, Hung AM, O’Donnell CJ, Edwards TL, Psaty BM, Tzoulaki I, Barnes MR, Wain LV, Elliott P, Caulfield MJ. Genetic analysis of over 1 million people identifies 535 new loci associated with blood pressure traits. Nat Genet. 2018;50(10):1412–25. Epub 20180917. doi: 10.1038/s41588-018-0205-x. PubMed PMID: 30224653; PMCID: PMC6284793.

9. Visscher PM, Wray NR, Zhang Q, Sklar P, McCarthy MI, Brown MA, Yang J. 10 Years of GWAS Discovery: Biology, Function, and Translation. Am J Hum Genet. 2017;101(1):5–22. doi: 10.1016/j.ajhg.2017.06.005. PubMed PMID: 28686856; PMCID: PMC5501872.

10. Hindorff LA, Sethupathy P, Junkins HA, Ramos EM, Mehta JP, Collins FS, Manolio TA. Potential etiologic and functional implications of genome-wide association loci for human diseases and traits. Proc Natl Acad Sci U S A. 2009;106(23):9362–7. Epub 20090527. doi: 10.1073/pnas.0903103106. PubMed PMID: 19474294; PMCID: PMC2687147.

11. Zhu Z, Zhang F, Hu H, Bakshi A, Robinson MR, Powell JE, Montgomery GW, Goddard ME, Wray NR, Visscher PM, Yang J. Integration of summary data from GWAS and eQTL studies predicts complex trait gene targets. Nat Genet. 2016;48(5):481–7. Epub 20160328. doi: 10.1038/ng.3538. PubMed PMID: 27019110.

12. Abbas M, Martin P, Lindsey ML, Bennett ES, Brown TL, Nzerue C, Williams CR, Gaye A. Intersecting transcriptomic landscapes of hypertension and kidney function in African American women. Am J Physiol Renal Physiol. 2025;329(1):F59–F70. Epub 20250530. doi: 10.1152/ajprenal.00067.2025. PubMed PMID: 40445960; PMCID: PMC12316382.

13. Consortium GT. The Genotype-Tissue Expression (GTEx) project. Nat Genet. 2013;45(6):580–5. doi: 10.1038/ng.2653. PubMed PMID: 23715323; PMCID: PMC4010069.

14. Robinson MD, McCarthy DJ, Smyth GK. edgeR: a Bioconductor package for differential expression analysis of digital gene expression data. Bioinformatics. 2010;26(1):139–40. Epub 20091111. doi: 10.1093/bioinformatics/btp616. PubMed PMID: 19910308; PMCID: PMC2796818.

15. Rus H, Cudrici C, Niculescu F. The role of the complement system in innate immunity. Immunol Res. 2005;33(2):103–12. doi: 10.1385/IR:33:2:103. PubMed PMID: 16234578.

16. Regal JF, Burwick RM, Fleming SD. The Complement System and Preeclampsia. Curr Hypertens Rep. 2017;19(11):87. Epub 20171018. doi: 10.1007/s11906-017-0784-4. PubMed PMID: 29046976; PMCID: PMC5849056.

17. Lynch AM, Eckel RH, Murphy JR, Gibbs RS, West NA, Giclas PC, Salmon JE, Holers VM. Prepregnancy obesity and complement system activation in early pregnancy and the subsequent development of preeclampsia. Am J Obstet Gynecol. 2012;206(5):428 e1-8. Epub 20120307. doi: 10.1016/j.ajog.2012.02.035. PubMed PMID: 22542119; PMCID: PMC10761005.

18. Ruan CC, Gao PJ. Role of Complement-Related Inflammation and Vascular Dysfunction in Hypertension. Hypertension. 2019;73(5):965–71. doi: 10.1161/HYPERTENSIONAHA.118.11210. PubMed PMID: 30929519.

19. Choo SY. The HLA system: genetics, immunology, clinical testing, and clinical implications. Yonsei Med J. 2007;48(1):11–23. doi: 10.3349/ymj.2007.48.1.11. PubMed PMID: 17326240; PMCID: PMC2628004.

20. Aisagbonhi O, Morris GP. Human Leukocyte Antigens in Pregnancy and Preeclampsia. Front Genet. 2022;13:884275. Epub 20220427. doi: 10.3389/fgene.2022.884275. PubMed PMID: 35571013; PMCID: PMC9093604.

21. Hiby SE, Walker JJ, O’Shaughnessy K M, Redman CW, Carrington M, Trowsdale J, Moffett A. Combinations of maternal KIR and fetal HLA-C genes influence the risk of preeclampsia and reproductive success. J Exp Med. 2004;200(8):957–65. Epub 20041011. doi: 10.1084/jem.20041214. PubMed PMID: 15477349; PMCID: PMC2211839.

22. Papuchova H, Meissner TB, Li Q, Strominger JL, Tilburgs T. The Dual Role of HLA-C in Tolerance and Immunity at the Maternal-Fetal Interface. Front Immunol. 2019;10:2730. Epub 20191209. doi: 10.3389/fimmu.2019.02730. PubMed PMID: 31921098; PMCID: PMC6913657.

23. Dunk CE, Bucher M, Zhang J, Hayder H, Geraghty DE, Lye SJ, Myatt L, Hackmon R. Human leukocyte antigen HLA-C, HLA-G, HLA-F, and HLA-E placental profiles are altered in early severe preeclampsia and preterm birth with chorioamnionitis. Am J Obstet Gynecol. 2022;227(4):641 e1- e13. Epub 20220719. doi: 10.1016/j.ajog.2022.07.021. PubMed PMID: 35863458.

24. Remmers EF, Cosan F, Kirino Y, Ombrello MJ, Abaci N, Satorius C, Le JM, Yang B, Korman BD, Cakiris A, Aglar O, Emrence Z, Azakli H, Ustek D, Tugal-Tutkun I, Akman-Demir G, Chen W, Amos CI, Dizon MB, Kose AA, Azizlerli G, Erer B, Brand OJ, Kaklamani VG, Kaklamanis P, Ben-Chetrit E, Stanford M, Fortune F, Ghabra M, Ollier WE, Cho YH, Bang D, O’Shea J, Wallace GR, Gadina M, Kastner DL, Gul A. Genome-wide association study identifies variants in the MHC class I, IL10, and IL23R-IL12RB2 regions associated with Behcet’s disease. Nat Genet. 2010;42(8):698–702. Epub 20100711. doi: 10.1038/ng.625. PubMed PMID: 20622878; PMCID: PMC2923807.

25. Kutllovci Hasani K, Ajeti N, Goswami N. Understanding Preeclampsia: Cardiovascular Pathophysiology, Histopathological Insights and Molecular Biomarkers. Med Sci (Basel). 2025;13(3). Epub 20250825. doi: 10.3390/medsci13030154. PubMed PMID: 40981151; PMCID: PMC12452302.

26. Deer E, Herrock O, Campbell N, Cornelius D, Fitzgerald S, Amaral LM, LaMarca B. The role of immune cells and mediators in preeclampsia. Nat Rev Nephrol. 2023;19(4):257–70. Epub 20230112. doi: 10.1038/s41581-022-00670-0. PubMed PMID: 36635411; PMCID: PMC10038936.

27. Wenzel UO, Kemper C, Bode M. The role of complement in arterial hypertension and hypertensive end organ damage. Br J Pharmacol. 2021;178(14):2849–62. Epub 20200819. doi: 10.1111/bph.15171. PubMed PMID: 32585035; PMCID: PMC10725187.

28. Madhur MS, Elijovich F, Alexander MR, Pitzer A, Ishimwe J, Van Beusecum JP, Patrick DM, Smart CD, Kleyman TR, Kingery J, Peck RN, Laffer CL, Kirabo A. Hypertension: Do Inflammation and Immunity Hold the Key to Solving this Epidemic? Circ Res. 2021;128(7):908–33. Epub 20210401. doi: 10.1161/CIRCRESAHA.121.318052. PubMed PMID: 33793336; PMCID: PMC8023750.

29. Singh MV, Chapleau MW, Harwani SC, Abboud FM. The immune system and hypertension. Immunol Res. 2014;59(1-3):243–53. doi: 10.1007/s12026-014-8548-6. PubMed PMID: 24847766; PMCID: PMC4313884.

30. Harmon AC, Cornelius DC, Amaral LM, Faulkner JL, Cunningham MW, Jr., Wallace K, LaMarca B. The role of inflammation in the pathology of preeclampsia. Clin Sci (Lond). 2016;130(6):409–19. doi: 10.1042/CS20150702. PubMed PMID: 26846579; PMCID: PMC5484393.

31. Campbell N, Robbins M, Nembaware H, Deer E, Cornelius D, LaMarca B. Immune Cells in Preeclampsia. Int J Mol Sci. 2025;27(1). Epub 20251221. doi: 10.3390/ijms27010074. PubMed PMID: 41515952; PMCID: PMC12785759.

32. Burwick RM, Feinberg BB. Eculizumab for the treatment of preeclampsia/HELLP syndrome. Placenta. 2013;34(2):201–3. Epub 20121208. doi: 10.1016/j.placenta.2012.11.014. PubMed PMID: 23228435.

33. Rodriguez-Iturbe B, Pons H, Johnson RJ. Role of the Immune System in Hypertension. Physiol Rev. 2017;97(3):1127–64. doi: 10.1152/physrev.00031.2016. PubMed PMID: 28566539; PMCID: PMC6151499.

34. Wu Y, Yu G, Jin K, Qian J. Advancing non-small cell lung cancer treatment: the power of combination immunotherapies. Front Immunol. 2024;15:1349502. Epub 20240702. doi: 10.3389/fimmu.2024.1349502. PubMed PMID: 39015563; PMCID: PMC11250065.

